# Childhood Anemia and Maternal Media Exposure in Sub-Saharan Africa: A Multi-Country Cross-Sectional Analysis Across Ghana, Nigeria, and Tanzania

**DOI:** 10.64898/2026.09.08.26362530

**Authors:** Enock Adu Bonsu, Daniel Ebo, Dorcas Doku

## Abstract

**Objectives:** To examine the association between maternal media exposure and childhood anemia in three sub-Saharan African countries, and to develop a two-stage multiple imputation approach for substantial hemoglobin outcome missingness in Demographic and Health Survey (DHS) data.

**Design:** Multi-country cross-sectional analysis of nationally representative DHS data.

**Setting:** Community-based household surveys in Ghana (2022), Nigeria (2023–24), and Tanzania (2022).

**Participants:** 42,944 children aged 6–59 months across 2,621 clusters.

**Primary and secondary outcome measures:** The primary outcome was anemia (altitude-adjusted hemoglobin <11.0 g/dL, WHO criteria); the primary exposure was a composite maternal media score (0–4 channels). Given 61.5% hemoglobin missingness, we imputed altitude-adjusted hemoglobin using multilevel Gaussian models and derived anemia deterministically, then estimated associations using population-averaged generalized estimating equations (pre-specified primary analysis), with six sensitivity analyses and exploratory mediation analysis.

**Results:** Media exposure was significantly associated with reduced anemia odds (OR 0.946, 95% CI 0.917 to 0.976, p<0.001; FMI 32.5%), a 5.4% reduction per additional weekly channel. Estimates were consistent across six sensitivity analyses (OR range 0.914–0.961), with no heterogeneity by country (I²=0%, p=0.482). Media exposure predicted antenatal care attendance and iron supplementation (both p<0.001), but neither mediated the anemia association.

**Conclusions:** Maternal media exposure was consistently associated with reduced childhood anemia across three sub-Saharan African settings. A two-stage multiple imputation approach addressed substantial (61.5%) outcome missingness while maintaining stable estimates, offering a transferable framework for DHS-based analyses. Prospective and intervention studies are needed before media-based communication can be recommended as a complementary anemia-control strategy.

**Strengths and Limitations of This Study:**

- This is among the first multi-country analyses to directly examine maternal media exposure and childhood anemia using nationally representative biomarker data from three sub-Saharan African countries.
- A validated two-stage multiple imputation approach addressed substantial (61.5%) hemoglobin missingness, with robustness confirmed across six pre-specified sensitivity analyses.
- The cross-sectional design precludes establishing temporality and cannot rule out reverse causation or residual confounding from unmeasured household factors.
- Maternal media exposure was measured as frequency of channel use only, without capturing content, quality, or health-specific programming.
- Mediation analysis relied on a single imputed dataset with non-clustered models, limiting direct comparability with the primary GEE-based estimate.

## Background and Introduction

Childhood anemia is a pressing public health challenge, especially in developing countries in sub-Saharan Africa, where over half of children under the age of five are affected [1,2]. Aside from immediate health impacts, anemia has negative implications on cognitive development, future economic productivity, educational attainment, and poverty cycles [1,3]. Although decades of interventions have sought to reduce the prevalence of childhood anemia, progress remains relatively slow, as several countries are continually faced with high prevalence rates [1,4].

Childhood anemia is attributed to multiple and complex factors [5,6]. Globally, the most common risk factor for anemia has been recognized as iron deficiency [3,7], however in sub-Saharan Africa, a combination of factors including nutritional challenges, infectious diseases such as malaria, genetic hemoglobinopathies, as well as chronic inflammation remain notable [3,7,8]. The implication of this multiplicity of risk factors is that efforts aimed at curbing child anemia in such regions ought to be multifaceted, taking into consideration both biomedical and socio-demographic factors [7,9]. Traditional approaches, centered on nutritional and biomedical intervention, remain constrained by poor coverage and adherence tied to education, health infrastructure, and poverty [2,3,6,10,11]. Household-level factors, particularly maternal knowledge and health behaviors, are increasingly recognized as critical determinants of child health outcomes, including anemia risk [2,3,9,11,12].

Mothers and primary caregivers, who make daily decisions about children’s nutrition and healthcare, rely heavily on access to information to make these decisions, especially where formal healthcare access is limited [13]. Mass media is a plausible and understudied channel for this information [14]. Prior research has established that maternal media exposure is associated with improved maternal healthcare-seeking behavior in sub-Saharan Africa, including antenatal care attendance and preventive health check-ups [15,16,17], and with improvements in some child health outcomes, such as growth stunting, operating in part through household health behaviors [14]. However, whether this association extends specifically to childhood anemia has not been directly examined.

This leaves two distinct gaps. First, no multi-country study has directly examined the association between maternal media exposure and childhood anemia using nationally representative biomarker data, despite established links between media exposure and related maternal health-seeking behaviors. Second, a substantial methodological barrier has likely contributed to this gap: hemoglobin measurement in DHS biomarker surveys is typically missing for 30–70% of sampled children, due to logistical constraints, child absence, and caregiver refusal. Most existing studies handling DHS biomarker data either exclude children with missing hemoglobin (complete-case analysis) or do not adequately address the missingness, both of which risk selection bias when the assumption of missingness completely at random (MCAR) does not hold. Multiple imputation offers a principled alternative, but its application to outcome biomarker variables, rather than covariates alone, in multi-country DHS analyses remains limited, and its performance under this degree of missingness is not well established.

This study addresses both gaps. Using nationally representative DHS data from Ghana, Nigeria, and Tanzania, we had two aims: (1) to estimate the association between maternal media exposure and childhood anemia using rigorous, cluster-aware, population-averaged modeling, and (2) to develop and validate a two-stage multiple imputation approach, imputing hemoglobin directly and deriving anemia status deterministically, for addressing substantial outcome missingness in multi-country DHS biomarker data, assessing its stability through comprehensive sensitivity analyses. Together, these aims provide both a rigorous test of a specific substantive hypothesis and a transferable methodological approach applicable to the many other DHS-based analyses facing similar missing-data challenges.

## Methods

### Study Design and Data Sources

We analyzed Demographic and Health Surveys (DHS) data from Ghana (2022), Nigeria (2023– 24), and Tanzania (2022), selected to represent diverse contexts in childhood anemia prevalence, economic development, and malaria endemicity. DHS employs two-stage stratified cluster sampling: enumeration areas selected with probability proportional to size, followed by systematic household sampling within clusters [18]. Hemoglobin was measured via capillary blood using the HemoCue Hb 301 system. Survey protocols were approved by country-specific institutional review boards and ICF International’s Institutional Review Board. Data are publicly available at https://dhsprogram.com.

### Study Population

Our analytical sample comprised 42,944 children aged 6–59 months across Ghana (n=8,349), Nigeria (n=24,945), and Tanzania (n=9,650), spanning 2,621 clusters. Children under 6 months were excluded per WHO hemoglobin assessment guidelines [19]. Child age was derived from the birth-history variable (b19) rather than the age-at-biomarker-measurement variable (hw1); the latter is populated only for children who completed hemoglobin measurement and would otherwise introduce substantial, non-random missingness into age itself, a variable central to both the eligibility filter and the imputation model. All sampled children meeting the age criteria were included regardless of hemoglobin measurement completion, to enable comprehensive missing-data assessment.

### Outcome and Exposure Measures

The primary outcome was anemia, defined as altitude-adjusted hemoglobin <11.0 g/dL for children aged 6–59 months, per WHO criteria [19]. Anemia status was derived directly from altitude-adjusted hemoglobin using this threshold for both observed and (following imputation) previously missing children, ensuring identical classification criteria across both groups. Maternal media exposure (primary exposure) was assessed via four channels: newspaper/magazine reading, radio listening, television viewing, and internet use (each categorized as at least weekly vs. less frequently). A composite media exposure score (range: 0– 4) summed weekly exposure across channels. Covariates included child characteristics (age, sex, birth order, recent illness), maternal factors (age, education, body mass index), household characteristics (wealth quintile, size, water/sanitation), health behaviors (antenatal care visits, iron supplementation), and geographic factors (urban/rural residence, region, country).

## Statistical Analysis

All analyses used R version 4.3.1 (two-sided α=0.05) [20]. We employed a potential outcomes framework, estimating adjusted associations rather than causal effects given potential unmeasured confounding. The population-averaged GEE model with two-stage multiple imputation (described below) constituted the single pre-specified primary analysis for testing the association between maternal media exposure and childhood anemia. All other analyses, sensitivity analyses SA1–SA6, country-stratified models, and mediation analyses, were secondary and exploratory, intended to assess the robustness and potential mechanisms of the primary estimate rather than to serve as independent tests of the primary hypothesis.

### Missing Data Characterization

We assessed missingness completely at random (MCAR) using Little’s test. To evaluate the plausibility of a missing-at-random (MAR) assumption, we fit a logistic regression model predicting hemoglobin missingness from all observed covariates, including child, maternal, household, and geographic characteristics, and maternal media exposure.

### Primary Analysis: Two-Stage Multiple Imputation with Population-Averaged Models

We addressed missing data using two-stage multiple imputation ensuring internal consistency between hemoglobin values and derived anemia status.

**Stage 1** – Covariate Imputation: Missing covariates were imputed using multivariate imputation by chained equations (MICE) generating 100 datasets [21]. Binary variables used logistic regression; continuous variables used predictive mean matching. The imputation model included all analysis variables plus auxiliary variables to strengthen MAR plausibility.

**Stage 2** – Hemoglobin Imputation: For each imputed covariate dataset, we fit multilevel Gaussian models predicting altitude-adjusted hemoglobin from all covariates with cluster random effects, generated predicted values with residual error, and derived anemia status from imputed hemoglobin using the identical 11.0 g/dL threshold applied to observed cases. Imputing the altitude-adjusted hemoglobin variable, rather than raw hemoglobin, ensures that observed and imputed anemia status are classified on an identical basis, since DHS derives observed anemia status from altitude-adjusted hemoglobin.

Convergence was verified via trace plots showing stability by iteration 20. Between-imputation variability in anemia prevalence was calculated to assess stability.

Population-Averaged Models Using Generalized Estimating Equations (GEE): For each imputed dataset, we fit population-averaged logistic regression models using GEE with exchangeable working correlation structure to account for clustering within communities. GEE provides population-averaged effect estimates, the expected change in anemia prevalence if maternal media exposure were increased across a population, which are most relevant for public health policy and intervention planning [22]. Models were fitted using the geepack package in R [23].

Pooling: Point estimates and variance components were pooled across the 100 imputed datasets using Rubin’s rules [24]. Given the large number of imputations (m=100), pooled inference used a large-sample normal approximation rather than the Barnard-Rubin t-distribution correction: in this dataset the latter produced degenerate degrees of freedom (df≈1) when between-imputation variance was small relative to within-imputation variance, artificially widening confidence intervals via heavy-tailed corrections not appropriate at this scale of m. This large-m normal approximation is an established alternative [25]. Fraction of Missing Information (FMI) was calculated for all parameters; FMI <40% was pre-specified as indicating stable estimates.

We fitted sequential models: Model 1 (child age and sex), Model 2 (adding socioeconomic factors), Model 3 (adding media exposure, PRIMARY), and Model 4 (adding health behaviors).

## Sensitivity Analyses

We assessed robustness through six sensitivity analyses:

SA1 – Multilevel Models: Three-level logistic regression with random intercepts for households nested within clusters using glmmTMB, applied to the full multiply imputed data and pooled identically to the primary analysis.

SA2 – Survey-Weighted Complete-Case Analysis: Survey-weighted logistic regression on complete cases incorporating DHS sampling weights with cluster-robust standard errors.

SA3 – Weighted Multiple Imputation: Post-imputation weighting, applying survey weights during analysis of the multiply imputed datasets.

SA4 – Country-Stratified Models: Separate models for Ghana, Nigeria, and Tanzania, each using the full two-stage multiple imputation procedure pooled across all 100 datasets via Rubin’s rules, identical to the primary analysis framework, to test heterogeneity across diverse contexts.

SA5 – Alternative Outcome Specifications: Continuous hemoglobin (g/dL) as outcome using linear GEE models.

SA6 – Missing Not at Random (MNAR) Sensitivity: δ-adjustment shifting imputed altitude-adjusted hemoglobin by increments (δ ∈ {−1.0 to +1.0} SD) to identify tipping points where conclusions would reverse.

Sensitivity analyses SA1–SA6 and country-stratified models (SA4) were designed to evaluate the robustness of the primary effect estimate across alternative analytical choices, modeling framework, weighting, sample restriction, outcome specification, and missing-data assumptions, rather than as independent hypothesis tests of separate research questions. Accordingly, no correction for multiple comparisons was applied.

### Mechanistic Exploration: Mediation Analysis

To explore potential pathways linking maternal media exposure to childhood anemia, we conducted exploratory mediation analyses examining whether the association operates through increased maternal health-seeking behaviors [26]. We focused on two hypothesized mediators identified a priori from the literature: (1) adequate antenatal care attendance (≥4 visits), and (2) iron supplementation during pregnancy. Using the first imputed dataset to facilitate bootstrap procedures, we fit mediation models using the mediation package in R with parametric assumptions and 1,000 bootstrap simulations for confidence interval estimation. For each mediator, we estimated total effect (c path), exposure-mediator association (a path), mediator-outcome association (b path), direct effect (c′ path), and indirect effect (proportion mediated). Models adjusted for child age, sex, and birth order; maternal age and education; and household wealth index and country.

Because the mediation package’s bootstrap-based estimation of average causal mediation effects does not natively accommodate GEE-based clustered variance estimation or pooling across multiply imputed datasets, mediation models were fit as single-dataset (first imputed dataset only), non-clustered generalized linear models rather than the full GEE-with-MI framework used in the primary analysis. As a result, total and direct effect estimates from this analysis use a different variance structure than the primary model and are not directly comparable to it in statistical significance, despite yielding closely similar odds ratios. These estimates are therefore reported as odds ratios and 95% confidence intervals only, without p-values, to avoid implying an independent significance test of the primary association.

### Patient and Public Involvement

This study used anonymized, publicly available secondary data from the Demographic and Health Surveys (DHS) program. As such, patients and members of the public were not directly involved in the design, conduct, reporting, or dissemination plans of this research. The DHS data were collected by trained field teams following nationally approved protocols, with informed consent obtained from participants at the time of data collection by the respective country survey teams and ICF International.

## Ethics Approval

The original DHS surveys received ethical clearance from ICF International’s Institutional Review Board and from the relevant national ethics review committee in each country (Ghana, Nigeria, Tanzania), with written or verbal informed consent obtained from participants or their guardians at the time of data collection. This study is a secondary analysis of fully de-identified, publicly available survey data and did not involve any new contact with participants; no additional institutional ethical approval was required or sought for this secondary analysis, consistent with standard practice for anonymized DHS data use.

## Results

### Study Population

The analytical sample comprised 42,944 children aged 6–59 months across Ghana (n=8,349), Nigeria (n=24,945), and Tanzania (n=9,650), spanning 2,621 clusters (Table 1, Figure 1). Anemia prevalence among children with observed hemoglobin was 57.8% overall (Ghana 54.2%, Nigeria 58.0%, Tanzania 60.6%). Mean media exposure score was 0.78 (SD 1.00), highest in Ghana (1.15) and lowest in Nigeria (0.68). Maternal education, wealth, and health-behavior indicators varied substantially by country, consistent with known cross-national differences in this region.

**Figure 1.**
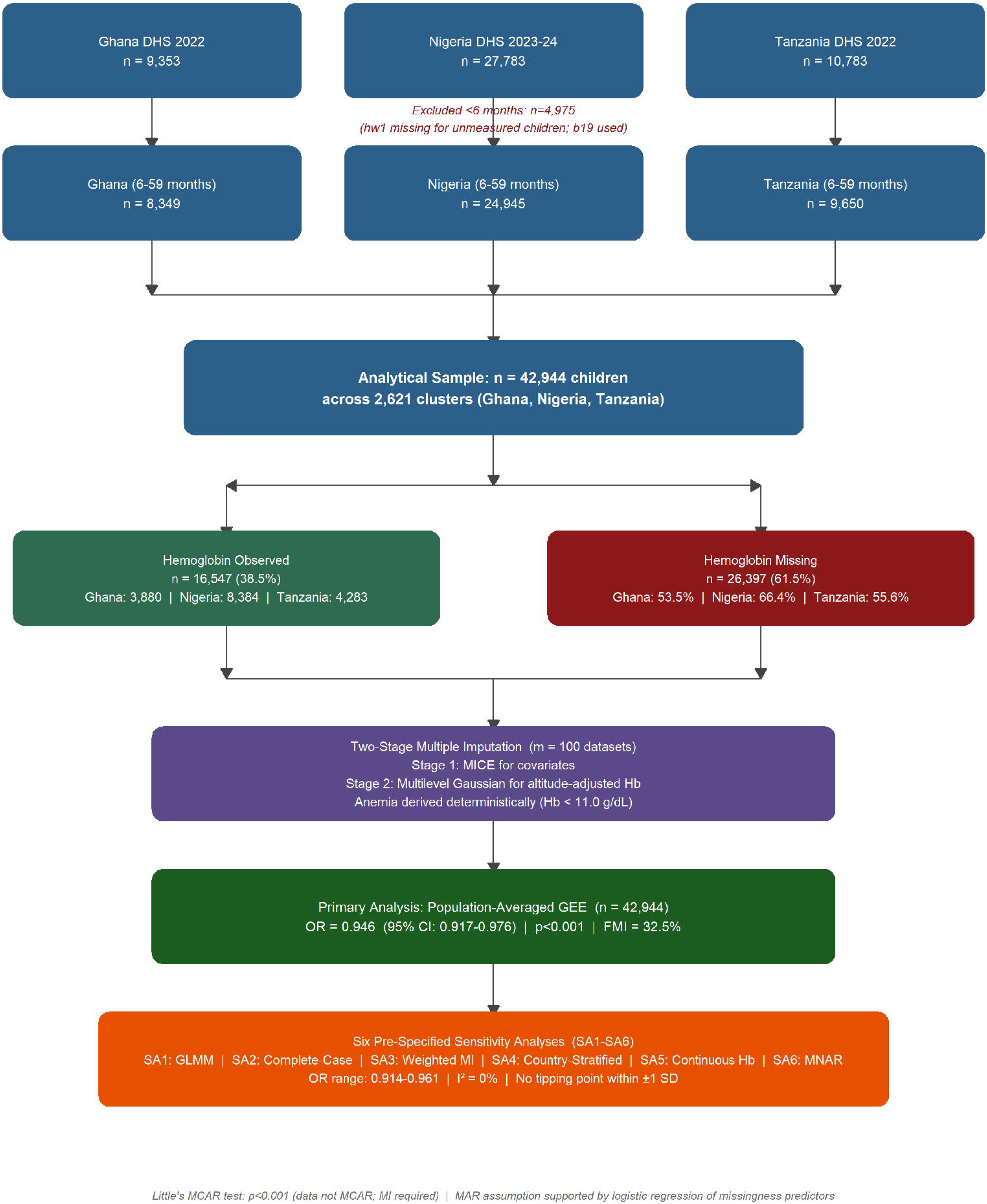
Study sample selection and analytical approach. Flow diagram showing derivation of the analytical sample from raw DHS enumeration in Ghana, Nigeria, and Tanzania, through the age-eligibility screen (6–59 months), hemoglobin observed/missing status, the two-stage multiple imputation procedure, the primary population-averaged GEE analysis, and the six pre-specified sensitivity analyses.

**Table 1.**
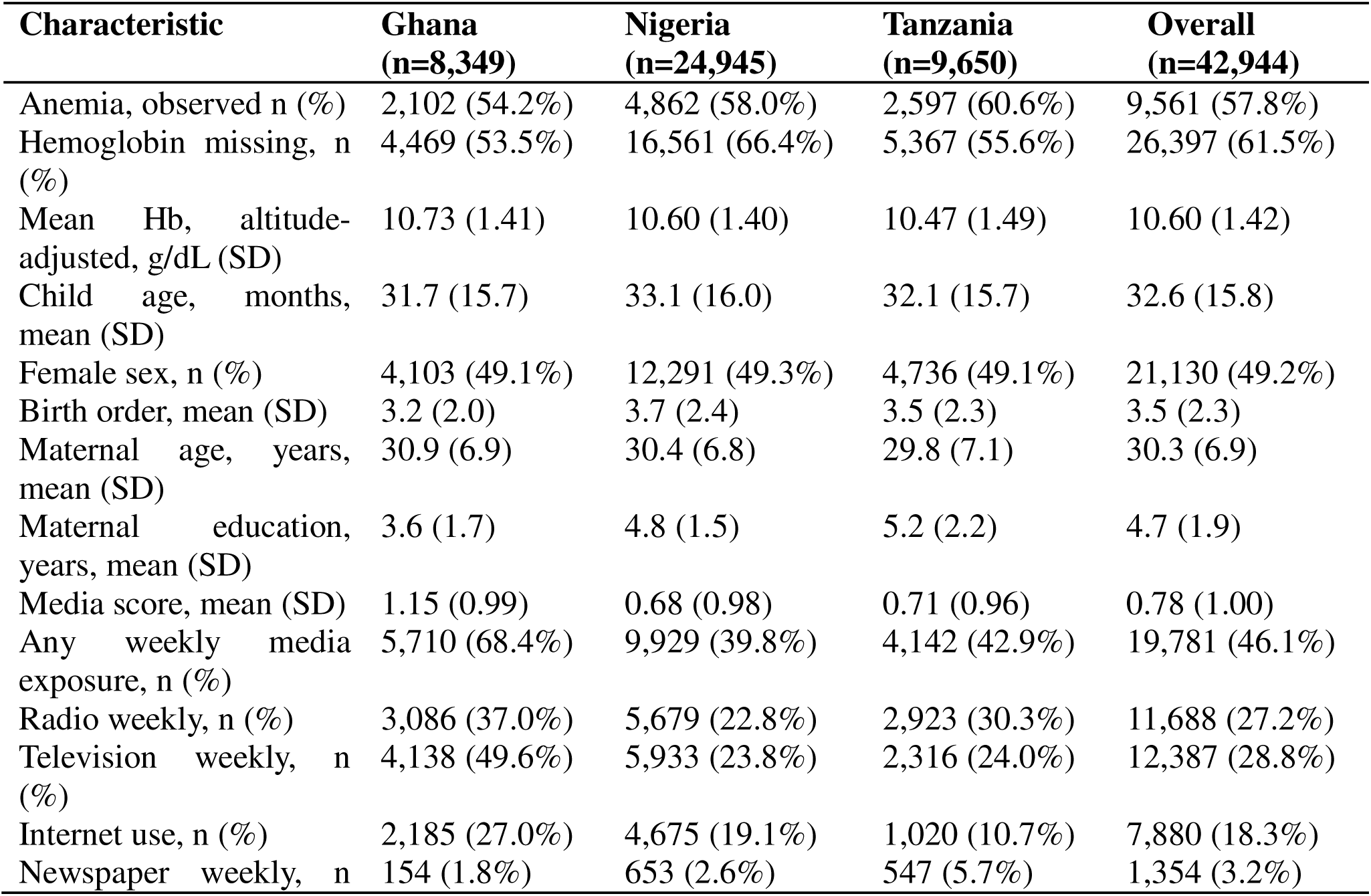
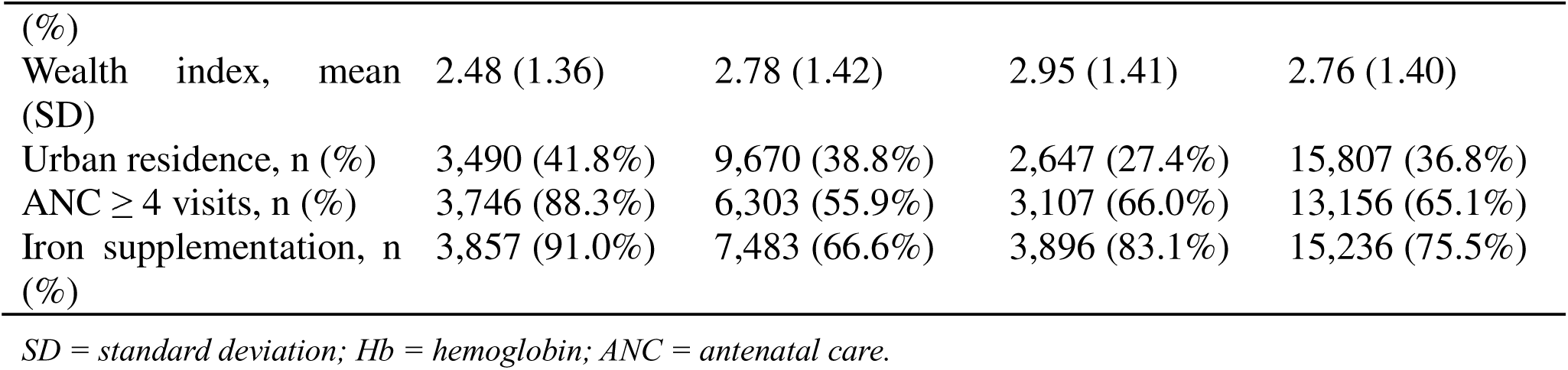
Baseline Characteristics by Country (n=42,944)

| Characteristic | Ghana<br>(n=8,349) | Nigeria<br>(n=24,945) | Tanzania<br>(n=9,650) | Overall<br>(n=42,944) |
| --- | --- | --- | --- | --- |
| Anemia, observed n (%) | 2,102 (54.2%) | 4,862 (58.0%) | 2,597 (60.6%) | 9,561 (57.8%) |
| Hemoglobin missing, n (%) | 4,469 (53.5%) | 16,561 (66.4%) | 5,367 (55.6%) | 26,397 (61.5%) |
| Mean Hb, altitude-adjusted, g/dL (SD) | 10.73 (1.41) | 10.60 (1.40) | 10.47 (1.49) | 10.60 (1.42) |
| Child age, months, mean (SD) | 31.7 (15.7) | 33.1 (16.0) | 32.1 (15.7) | 32.6 (15.8) |
| Female sex, n (%) | 4,103 (49.1%) | 12,291 (49.3%) | 4,736 (49.1%) | 21,130 (49.2%) |
| Birth order, mean (SD) | 3.2 (2.0) | 3.7 (2.4) | 3.5 (2.3) | 3.5 (2.3) |
| Maternal age, years, mean (SD) | 30.9 (6.9) | 30.4 (6.8) | 29.8 (7.1) | 30.3 (6.9) |
| Maternal education, years, mean (SD) | 3.6 (1.7) | 4.8 (1.5) | 5.2 (2.2) | 4.7 (1.9) |
| Media score, mean (SD) | 1.15 (0.99) | 0.68 (0.98) | 0.71 (0.96) | 0.78 (1.00) |
| Any weekly media exposure, n (%) | 5,710 (68.4%) | 9,929 (39.8%) | 4,142 (42.9%) | 19,781 (46.1%) |
| Radio weekly, n (%) | 3,086 (37.0%) | 5,679 (22.8%) | 2,923 (30.3%) | 11,688 (27.2%) |
| Television weekly, n (%) | 4,138 (49.6%) | 5,933 (23.8%) | 2,316 (24.0%) | 12,387 (28.8%) |
| Internet use, n (%) | 2,185 (27.0%) | 4,675 (19.1%) | 1,020 (10.7%) | 7,880 (18.3%) |
| Newspaper weekly, n | 154 (1.8%) | 653 (2.6%) | 547 (5.7%) | 1,354 (3.2%) |

|  |  |  |  |  |  |
| --- | --- | --- | --- | --- | --- |
| (%) |  |  |  |  |  |
| Wealth index, mean (SD) | 2.48 (1.36) | 2.78 (1.42) | 2.95 (1.41) | 2.76 (1.40) |  |
| Urban residence, n (%) | 3,490 (41.8%) | 9,670 (38.8%) | 2,647 (27.4%) | 15,807 (36.8%) |  |
| ANC ≥ 4 visits, n (%) | 3,746 (88.3%) | 6,303 (55.9%) | 3,107 (66.0%) | 13,156 (65.1%) | |
| Iron supplementation, n (%) | 3,857 (91.0%) | 7,483 (66.6%) | 3,896 (83.1%) | 15,236 (75.5%) |  |

*SD = standard deviation; Hb = hemoglobin; ANC = antenatal care.*

### Missing Data Patterns

Hemoglobin measurements were missing for 26,397 children (61.5%) (Table 2, Part A). Missingness was not completely at random (Little’s MCAR test: χ²=42,983.3, df=119, p<0.001). Rather than being concentrated in one dominant predictor, missingness was jointly associated with several observed covariates (Table 2, Part B): country showed the strongest signal, particularly higher missingness in Nigeria; wealth index, maternal age, and, to a modest degree, child age were also significant predictors (child age OR=1.007 per month; over the full 6–59 month range this corresponds to roughly a 40–45% relative shift in odds of missingness, notable but not dominant). Maternal media exposure itself was unrelated to missingness (p=0.427), supporting the plausibility of MAR conditional on observed covariates for the exposure of central interest.

**Table 2.** Missing Data Patterns and Predictors Part A: Missingness of Hemoglobin by Country. Part B: Predictors of Hemoglobin Missingness

Part A: Missingness of Hemoglobin by Country
| Country | Total N | Hb Observed | Hb Missing | % Missing | Anemia Prev.* (%) | Mean Hb (g/dL)* |
| --- | --- | --- | --- | --- | --- | --- |
| Ghana | 8,349 | 3,880 | 4,469 | 53.5 | 54.2 | 10.7 |
| Nigeria | 24,945 | 8,384 | 16,561 | 66.4 | 58.0 | 10.6 |
| Tanzania | 9,650 | 4,283 | 5,367 | 55.6 | 60.6 | 10.7 |
| Overall | 42,944 | 16,547 | 26,397 | 61.5 | 57.8 | 10.6 |

\*Among children with observed hemoglobin only.

Part B: Predictors of Hemoglobin Missingness
| Predictor | Odds Ratio | 95% CI | p-value |
| --- | --- | --- | --- |
| Country: Nigeria (vs. Ghana) | 1.456 | 1.360–1.560 | <0.001 |
| Country: Tanzania (vs. Ghana) | 1.118 | 1.035–1.208 | 0.004 |
| Child age (per month) | 1.007 | 1.005–1.008 | <0.001 |
| Female child | 0.977 | 0.931–1.025 | 0.341 |
| Birth order | 1.011 | 0.994–1.029 | 0.199 |
| Maternal age (per year) | 0.990 | 0.985–0.996 | <0.001 |
| Maternal education (per year) | 0.990 | 0.976–1.004 | 0.146 |
| Wealth index | 0.954 | 0.931–0.978 | <0.001 |
| Urban residence | 1.056 | 0.996–1.119 | 0.068 |
| Media score | 1.011 | 0.984–1.039 | 0.427 |

Logistic regression predicting probability of missing hemoglobin, adjusted for all covariates shown. Little's MCAR test: $\chi^2=42,983.3$ , $df=119$ , $p<0.001$ .

### Primary Analysis: Population-Averaged Multiple Imputation Results

Using population-averaged GEE models with two-stage multiple imputation across 100 datasets, maternal media exposure was significantly associated with reduced odds of childhood anemia (OR=0.946, 95% CI 0.917–0.976, p<0.001) (Table 3). This corresponds to a 5.4% reduction in anemia odds per additional weekly media channel. The fraction of missing information (FMI) for media score was 32.5%, below the pre-specified 40% stability threshold. Child age (OR=0.974 per month, p<0.001), female sex (OR=0.850 vs. male, p<0.001), and wealth index (OR=0.892, p<0.001) were independently protective; birth order (OR=1.055, p<0.001) and residence in Nigeria (OR=1.284) or Tanzania (OR=1.442, both vs. Ghana, p<0.001) were associated with higher odds of anemia. All FMI values were below 40% except maternal education (52.5%, moderate stability). Imputed hemoglobin distributions closely tracked observed values across countries and age groups (Supplementary figure S1).

**Table 3.** Primary Analysis: Population-Averaged GEE with Multiple Imputation (n=42,944)

| Variable | OR | 95% CI | p-value | FMI (%) |
| --- | --- | --- | --- | --- |
| Media score | 0.946 | 0.917–0.976 | <0.001 | 32.5 |
| Child age (months) | 0.974 | 0.973–0.976 | <0.001 | 37.1 |
| Female sex (ref: male) | 0.850 | 0.810–0.891 | <0.001 | 32.0 |
| Birth order | 1.055 | 1.038–1.073 | <0.001 | 35.7 |
| Maternal age | 0.982 | 0.977–0.988 | <0.001 | 34.7 |
| Maternal education (years) | 0.987 | 0.971–1.003 | 0.123 | 52.5 |
| Wealth index | 0.892 | 0.867–0.919 | <0.001 | 35.2 |
| Urban residence | 0.962 | 0.886–1.045 | 0.356 | 33.3 |
| Country: Nigeria (ref: Ghana) | 1.284 | 1.176–1.400 | <0.001 | 23.3 |
| Country: Tanzania (ref: Ghana) | 1.442 | 1.297–1.602 | <0.001 | 25.9 |
| (Intercept) | 6.605 | 5.554–7.856 | <0.001 | 38.1 |

Population-averaged estimates from GEE models with exchangeable correlation structure. Results pooled across 100 multiply imputed datasets using Rubin's rules with a large-sample normal approximation (see Methods). FMI = Fraction of Missing Information. OR = odds ratio; CI = confidence interval.

## Sensitivity Analyses

Effect estimates for media exposure were highly consistent across all six sensitivity analyses, ranging OR=0.914–0.961 (Table 4, Figure 2). GLMM cluster-specific models (SA1) closely matched the primary estimate (OR=0.942, 95% CI 0.912–0.974, p<0.001). Survey-weighted complete-case analysis (SA2, n=11,272) showed a consistent, significant association (OR=0.946, 95% CI 0.895–0.999, p=0.047). Weighted multiple imputation (SA3) confirmed the estimate was not driven by sampling weights (OR=0.941, 95% CI 0.906–0.978, p=0.002). Country-stratified models using the full MI-pooling procedure (SA4) showed consistent protective point estimates in all three countries, Ghana (OR=0.914, 95% CI 0.852–0.979, p=0.010), Nigeria (OR=0.961, 95% CI 0.920–1.004, p=0.077), Tanzania (OR=0.947, 95% CI 0.888–1.009, p=0.090), with no evidence of heterogeneity (I²=0%, Q=1.461, p=0.482); individual country estimates did not all reach significance, consistent with reduced power at the country level rather than inconsistency of effect. Continuous hemoglobin outcome analysis (SA5) corroborated the direction (β=0.046 g/dL per channel, 95% CI 0.025–0.066, p<0.001). MNAR δ-adjustment (SA6) found no tipping point within ±1 SD (OR range 0.944–0.951 across the tested range), indicating the conclusion is robust to plausible departures from MAR (Supplementary figure S2).

**Figure 2.**
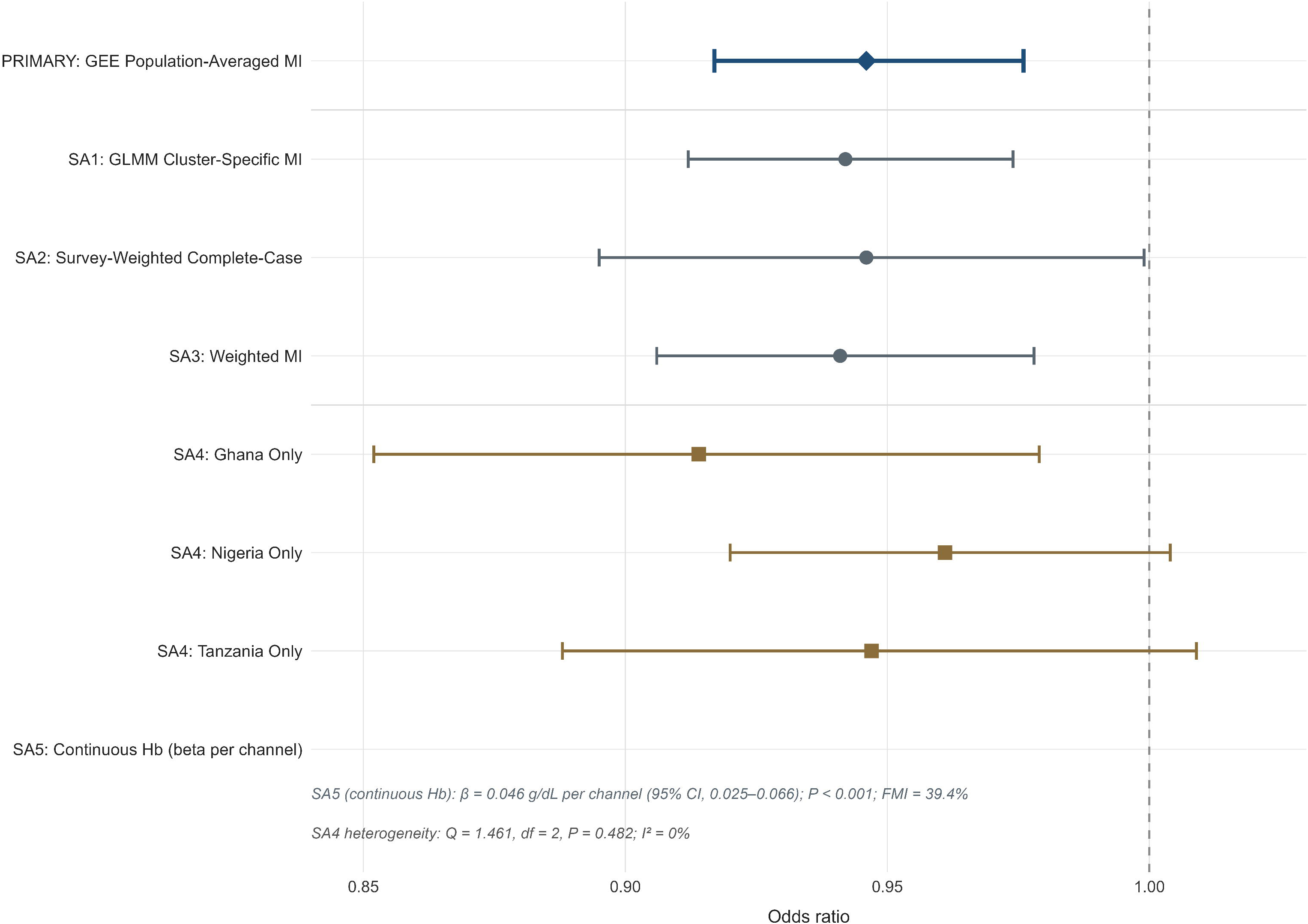
Forest plot of odds ratios for maternal media exposure and childhood anemia. Estimates are shown for the primary analysis and secondary sensitivity analyses (SA1–SA5), including country-stratified estimates for Ghana, Nigeria, and Tanzania under SA4. Points represent odds ratios; horizontal lines represent 95% confidence intervals; the dashed vertical line indicates the null value (OR=1.0).

**Table 4.** Sensitivity Analyses: Media Exposure and Childhood Anemia.

| Analysis | n | OR / $\beta$ (95% CI) | p-value | FMI |
| --- | --- | --- | --- | --- |
| Primary: GEE Population-Averaged MI | 42,944 | 0.946 (0.917–0.976) | <0.001 | 32.5% |
| SA1: GLMM Cluster-Specific MI | 42,944 | 0.942 (0.912–0.974) | <0.001 | 33.0% |
| SA2: Survey-Weighted Complete-Case | 11,272 | 0.946 (0.895–0.999) | 0.047 | — |
| SA3: Weighted MI | 42,944 | 0.941 (0.906–0.978) | 0.002 | 31.0% |
| SA4: Ghana | 8,349 | 0.914 (0.852–0.979) | 0.010 | 30.8% |
| SA4: Nigeria | 24,945 | 0.961 (0.920–1.004) | 0.077 | 38.2% |
| SA4: Tanzania | 9,650 | 0.947 (0.888–1.009) | 0.090 | 31.1% |
| SA4: Heterogeneity | — | $I^2=0\%$, $Q=1.461$, $df=2$ | 0.482 | — |
| SA5: Continuous Hemoglobin ($\beta$, g/dL) | 42,944 | 0.046 (0.025–0.066) | <0.001 | 39.4% |
| SA6: MNAR δ-Adjustment | 42,944 | No tipping point, $\pm 1$ SD | — | — |

*All analyses adjusted for child, maternal, household, and geographic factors, matching the primary model. MI = multiple imputation; GEE = generalized estimating equations; GLMM = generalized linear mixed models; FMI = fraction of missing information. SA1–SA6 are secondary/exploratory robustness checks on the single pre-specified primary estimate; no multiplicity correction was applied.*

## Mediation Analysis

Media exposure was strongly associated with both antenatal care attendance (≥4 visits: OR=1.322, 95% CI 1.263–1.385, p<0.001) and iron supplementation (OR=1.320, 95% CI 1.255– 1.390, p<0.001) (Table 5). Neither mediator was itself significantly associated with anemia (ANC: OR=1.046, 95% CI 0.973–1.124, p=0.223; iron: OR=0.945, 95% CI 0.875–1.020, p=0.147), and neither showed evidence of meaningful mediation (ACME 95% CIs spanned zero for both pathways). The total effect from this exploratory model (OR=0.956, 95% CI 0.933– 0.980) closely tracked the primary estimate (OR=0.946), and direct effects after adjusting for each mediator were essentially unchanged (OR=0.959 and 0.964, respectively), indicating the association does not operate primarily through these two specific healthcare-utilization pathways (Supplementary figure S3).

**Table 5.**
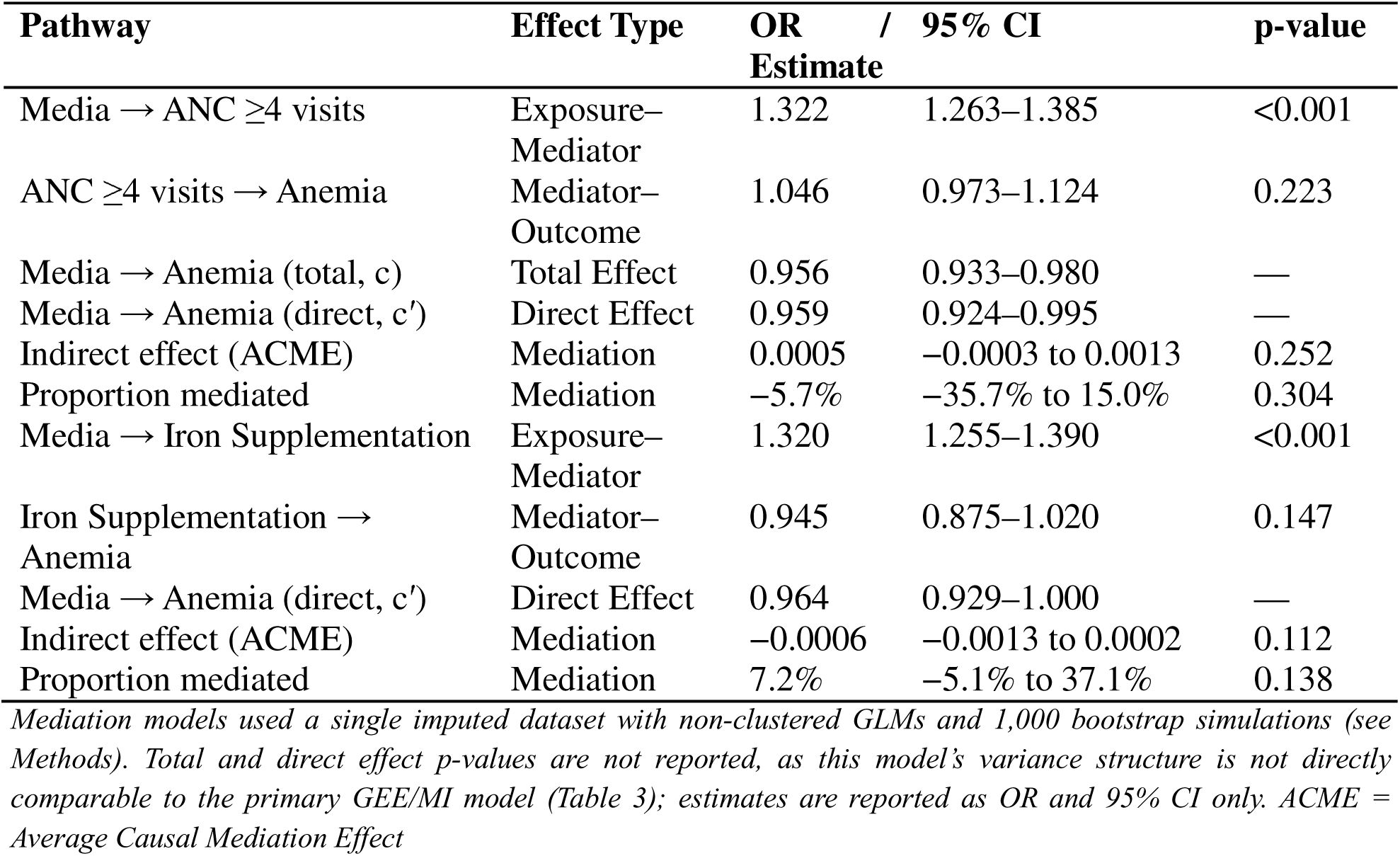
Mediation Analysis: Maternal Health Behaviors as Potential Pathways (first imputed dataset, n=42,944)

## Discussion Principal Findings

In this multi-country analysis of 42,944 children across Ghana, Nigeria, and Tanzania, maternal media exposure was associated with significantly reduced odds of childhood anemia (OR=0.946, 95% CI 0.917–0.976, p<0.001), an association that was highly consistent, in both direction and magnitude, across six sensitivity analyses varying in modeling framework, weighting, missing-data handling, and country. This consistency, combined with the absence of cross-country heterogeneity and a plausible (though not fully confirmed) mechanism via maternal health-behavior pathways, supports a genuine, modest, and generalizable association rather than an artifact of any single analytical choice.

### Interpretation of the Mediation Findings

Although media exposure strongly predicted two commonly hypothesized health-seeking behaviors, antenatal care attendance and iron supplementation, neither mediated the media-anemia association. This suggests the protective association, if causal, more likely operates through pathways not captured by these two specific measures, such as household-level nutrition and feeding practices, caregiver recognition of and response to childhood illness, or broader maternal health literacy and decision-making autonomy that jointly influence multiple behaviors without being reducible to formal healthcare utilization; media exposure has also been linked to stronger health information literacy and earlier illness recognition in comparable low-and middle-income settings [27,28,29]. These remain speculative given the data available here and warrant direct measurement in future work.

### Methodological Contributions

This study demonstrates that substantial (>60%) hemoglobin missingness in multi-country DHS data can be addressed via two-stage multiple imputation, imputing hemoglobin directly and deriving anemia deterministically, while retaining low fractions of missing information (<40% for nearly all parameters) and stable, well-calibrated pooled estimates. This addresses a recognized gap in DHS-based research, where analyses often either restrict to complete cases or acknowledge missingness without addressing it analytically, despite established guidance on missing data and multiple imputation for epidemiological research [30,31]. Two further points are worth highlighting for researchers working with similar data.

First, variable selection for imputation models matters more than it might appear. An early iteration of this analysis used the DHS variable recording age at biomarker measurement, which is populated only for children who completed the hemoglobin measurement, meaning it was missing for the majority of children whose hemoglobin also needed imputation, and created a spurious, artificially strong apparent relationship between child age and missingness status. Substituting the birth-history age variable, populated for all listed children, both corrected the imputation model’s information content and substantially changed the missingness picture: the true per-month association between age and missingness is modest and diffuse across several covariates (chiefly country and wealth), not the dominant single predictor an incorrectly specified variable had suggested. This is a specific, checkable pitfall relevant to any DHS-based analysis using measurement-time rather than birth-history age variables as imputation predictors.

Second, degrees-of-freedom handling in Rubin’s-rules pooling deserves explicit attention at large m. The standard finite-sample (Barnard-Rubin) correction can produce degenerate, near-unity degrees of freedom when between-imputation variance is small relative to within-imputation variance, a scenario more likely with well-specified imputation models and large m, leading to implausibly wide, heavy-tailed confidence intervals. A large-sample normal approximation avoided this problem here and is a defensible alternative under these conditions [25], but researchers should verify which regime applies to their own data rather than defaulting to either method without inspection.

Third, and related: an early country-stratified sensitivity analysis pooled across only a single imputed dataset rather than the full multiple-imputation procedure, understating uncertainty and producing individually significant country-level estimates that did not persist once pooled properly across all 100 imputations, though the direction and approximate magnitude were unaffected. Any secondary or subgroup analysis run alongside a multiply-imputed primary analysis should be pooled using the identical procedure, not a simplified single-dataset substitute, to avoid this class of error.

### Strengths and Limitations

Strengths include a large, multi-country, nationally representative sample; systematic characterization of missingness mechanisms; a single pre-specified primary analysis with clearly designated secondary analyses; and consistency of the effect estimate across modeling frameworks, weighting schemes, and countries.

Limitations include the cross-sectional design, which precludes establishing temporality or excluding residual confounding or reverse causation, for example, unmeasured household health engagement could plausibly influence both media consumption and child health outcomes. The composite media score captures frequency of channel use, not content or health-specific programming exposure. The MAR assumption, while supported by the missingness-predictor analysis and MNAR sensitivity analysis, cannot be definitively verified. Country-stratified estimates, while directionally and quantitatively consistent, did not all individually reach conventional significance, reflecting reduced power at the country level rather than inconsistency of effect; a formally powered multi-country design would strengthen confidence in the per-country estimates specifically. Finally, this analysis reflects a corrected re-analysis of prior work; while each correction (age variable, anemia derivation basis, pooling method) is independently justified and disclosed above, readers should weigh the overall estimate accordingly and treat the finding as warranting independent replication.

### Implications for Research and Practice

For DHS-based research, this study offers a validated approach to substantial biomarker outcome missingness and two specific, checkable pitfalls, imputation-predictor selection and subgroup pooling completeness, that likely affect other analyses using similar data and methods. For the substantive question, the consistency of a modest protective association across diverse contexts and analytical approaches supports further investigation of media-based nutrition and health communication as a potential complement to biomedical anemia-control efforts, though the observational, cross-sectional design means this should inform hypothesis generation for prospective or intervention-based research rather than direct policy translation on its own.

## Conclusion

In this multi-country analysis of 42,944 children, maternal media exposure was associated with a modest, statistically significant, and highly consistent reduction in the odds of childhood anemia across Ghana, Nigeria, and Tanzania (OR=0.946, 95% CI 0.917–0.976, p<0.001), robust across six sensitivity analyses varying in modeling approach, weighting, and missing-data handling. The association did not appear to operate through antenatal care attendance or iron supplementation specifically, suggesting alternative mechanisms warranting further study. Methodologically, this analysis demonstrates a validated two-stage multiple imputation approach for substantial DHS biomarker missingness, alongside two specific and generalizable lessons, on imputation-predictor selection and on complete pooling in subgroup analyses, relevant to the broader community of DHS-based researchers.

## Data Availability Statement

The data used in this study are publicly available from the Demographic and Health Surveys (DHS) Program. Researchers may request access to the Ghana 2022, Nigeria 2023-24, and Tanzania 2022 DHS datasets free of charge through the DHS Program website at https://dhsprogram.com. Registration and project approval are required prior to data access. The statistical analysis plan for this study is publicly registered on the Open Science Framework (OSF DOI: 10.17605/OSF.IO/HKA4F). All R code used for the analyses is publicly archived on Zenodo (https://doi.org/10.5281/zenodo.22666013).

## Funding Statement

This research received no specific grant from any funding agency in the public, commercial, or not-for-profit sectors.

## Competing Interests

None declared.

## Author Contributions

Enock Adu Bonsu conceptualized the study. Enock Adu Bonsu, Daniel Ebo, and Dorcas Doku designed the analytical framework and drafted the manuscript. Enock Adu Bonsu conducted all statistical analyses. Daniel Ebo and Dorcas Doku contributed to the interpretation of findings and provided critical revisions to the manuscript for important intellectual content. All authors approved the final version for submission.

## Acknowledgments

The authors thank the Demographic and Health Surveys (DHS) Program for making the Ghana, Nigeria, and Tanzania survey data publicly available. No persons other than the listed authors made a substantial contribution to this work.

## Notes

### Competing Interest Statement

The authors have declared no competing interest.

### Clinical Protocols

https://doi.org/10.17605/OSF.IO/HKA4F

### Author Declarations

The Institutional Review Board of ICF International gave ethical approval for this work through the original Demographic and Health Surveys data collection in Ghana (2022), Nigeria (2023-24), and Tanzania (2022). The National Ethics Committee of the Ghana Health Service gave ethical approval for the Ghana DHS 2022. The National Health Research Ethics Committee of Nigeria gave ethical approval for the Nigeria DHS 2023-24. The National Institute for Medical Research of Tanzania gave ethical approval for the Tanzania DHS 2022. The University of Arizona Institutional Review Board waived ethical approval for this work as it involves only secondary analysis of fully de-identified publicly available data.

