## Supplementary Figures and Materials for "Childhood Anemia and Maternal Media Exposure in Sub-Saharan Africa: A Multi-Country Cross-Sectional Analysis Across Ghana, Nigeria, and Tanzania"

**Supplemental Material**

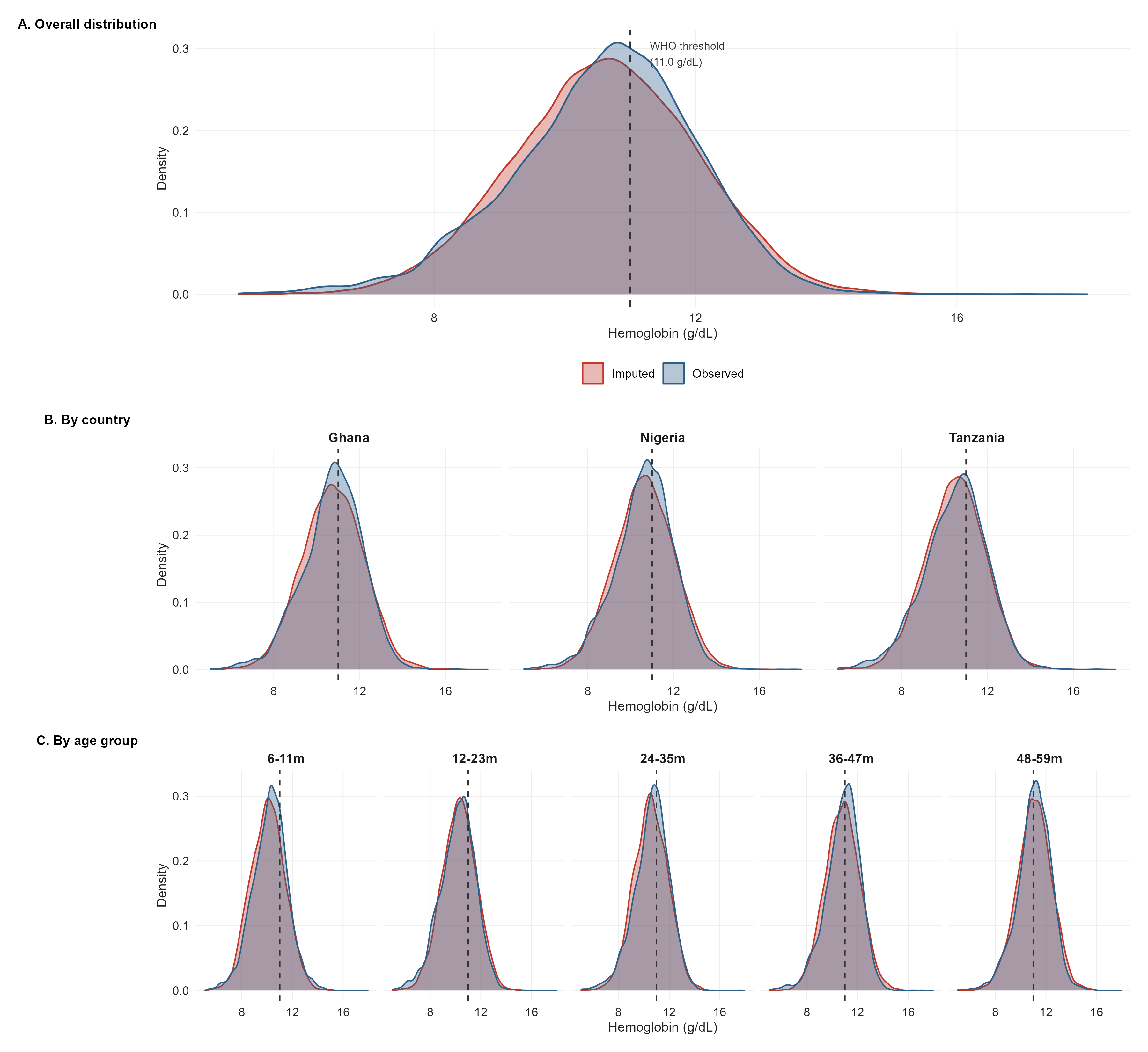

**Supplementary Figure 1.** Distribution of altitude-adjusted hemoglobin, observed versus imputed values (first imputed dataset). (A) Overall distribution; (B) by country; (C) by child age group. The dashed vertical line marks the WHO anemia threshold (11.0 g/dL). Imputed and observed distributions overlap closely across all subgroups, with no evidence of truncation or systematic shift, supporting the validity of the two-stage imputation approach described in Methods.

*Note: Imputed values shown are from the first of 100 imputed datasets, for illustration; validation statistics reported in Methods and Results (e.g., mean difference, between-imputation stability of anemia prevalence) are computed across all 100 datasets.*

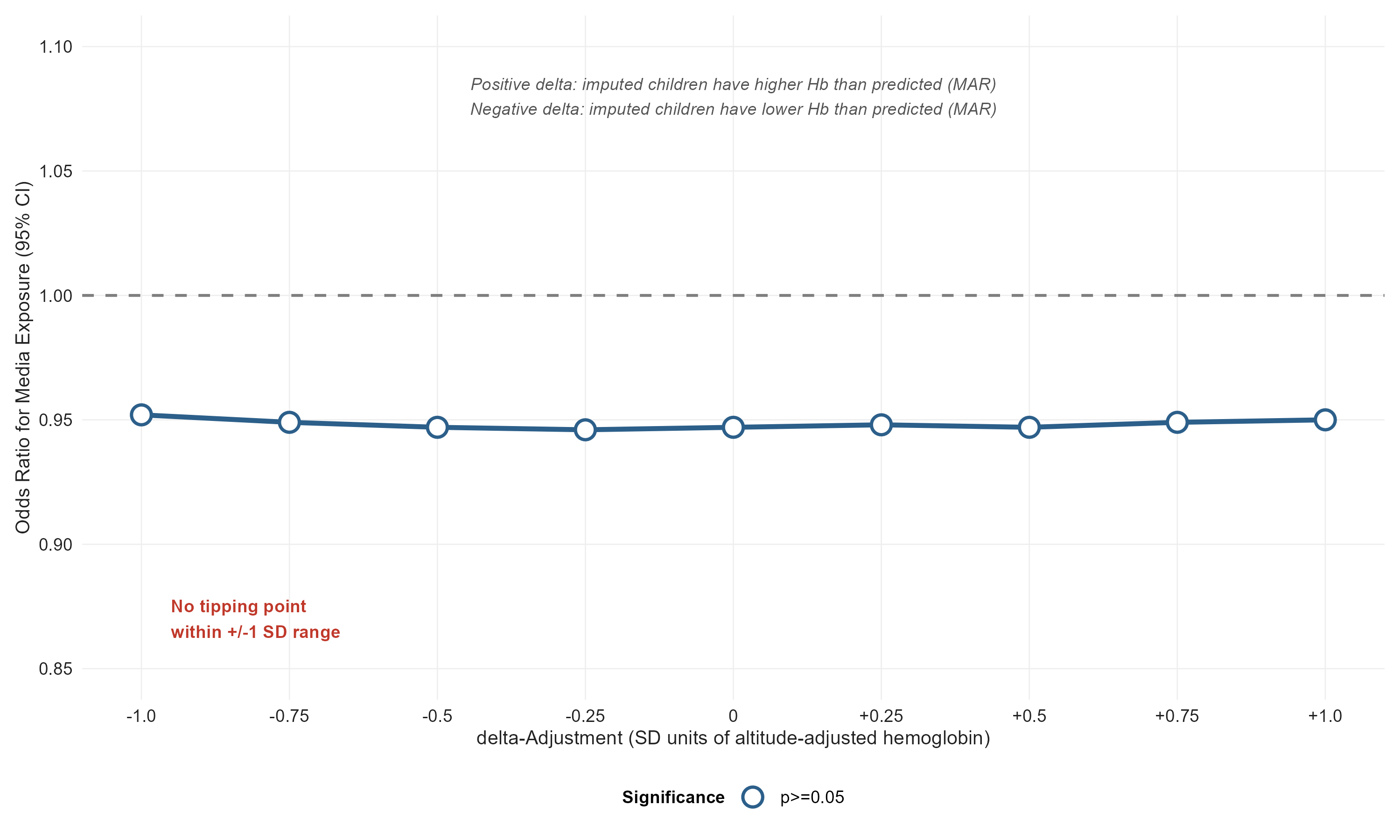

**Supplementary Figure 2.** Missing Not at Random (MNAR) δ-adjustment sensitivity analysis (SA6). Imputed altitude-adjusted hemoglobin values were shifted by δ = −1.0 to +1.0 SD to assess whether the primary conclusion is robust to departures from the missing-at-random (MAR) assumption. The odds ratio for maternal media exposure remained stable (0.944–0.951) and did not cross the null across the full tested range, indicating no tipping point within ±1 SD.

*Note: Corresponds to sensitivity analysis SA6 in Table 4. A positive δ corresponds to imputed children having systematically higher hemoglobin than predicted by the imputation model; a negative δ, systematically lower.*

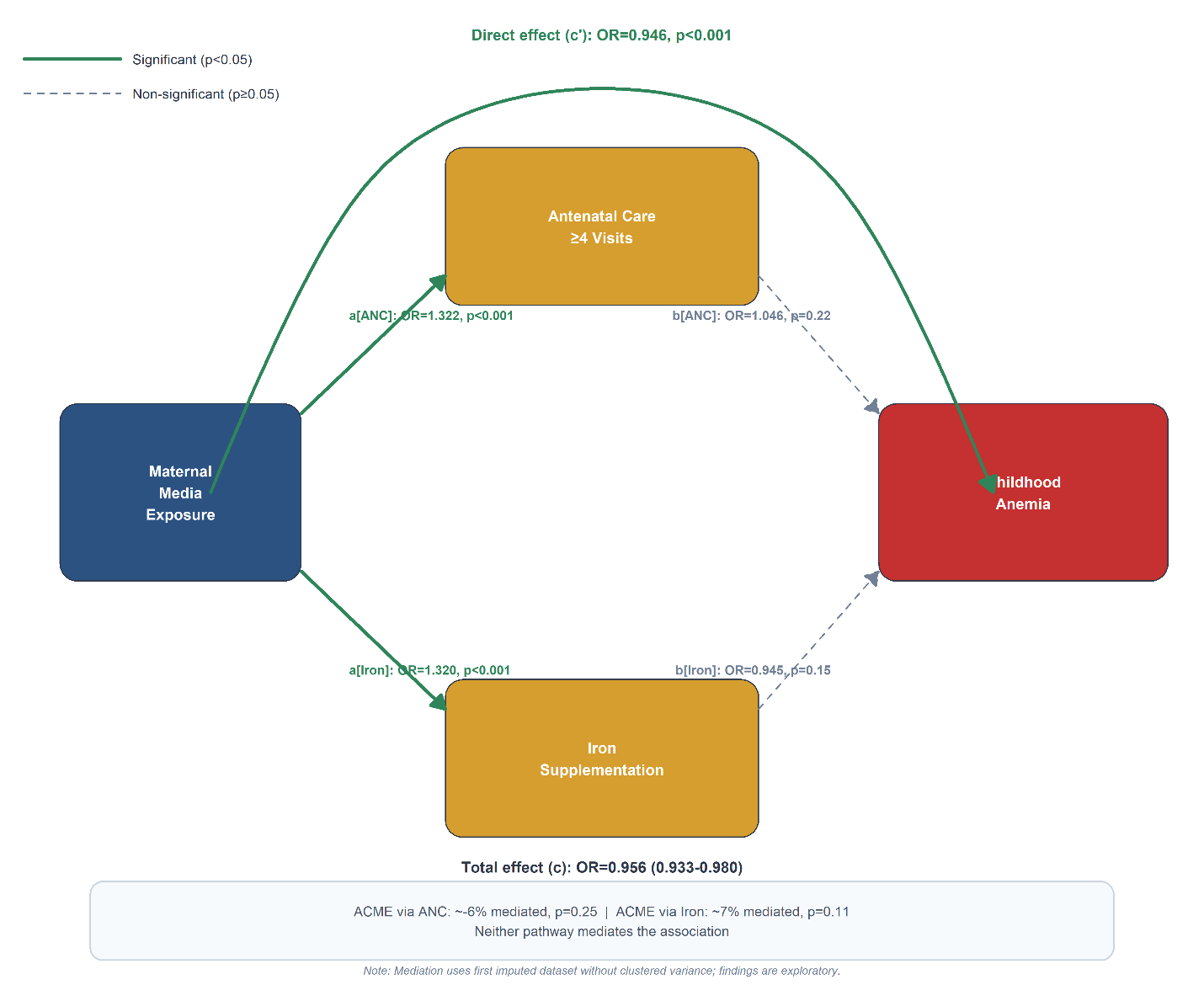

**Supplementary Figure 3.** Path diagram for the exploratory mediation analysis (Table 5). Solid green arrows indicate statistically significant paths (p<0.05); dashed grey arrows indicate non-significant paths. Media exposure was strongly associated with both antenatal care attendance (a[ANC]) and iron supplementation (a[Iron]), but neither mediator was itself associated with childhood anemia (b[ANC], b[Iron]), and the direct effect of media exposure on anemia (c′) remained essentially unchanged from the total effect (c), indicating minimal mediation through either pathway.

*Note: Mediation models used the first imputed dataset with non-clustered generalized linear models; total and direct effect estimates use a different variance structure than the primary GEE/multiple-imputation model and are not directly comparable to it in statistical significance (see Methods and Table 5 note).*
